# Detection of mycobacterial pulmonary diseases via breath analysis in clinical practice

**DOI:** 10.1101/2023.05.23.23290378

**Authors:** Biyi Su, Yong Feng, Haibin Chen, Jialou Zhu, Mengqi He, Lijuan Wu, Qing Sheng, Ping Guan, Pinru Chen, Haobin Kuang, Dexian Li, Weiyong Wang, Zhiyu Feng, Yigang Tan, Jianxiong Liu, Yaoju Tan

## Abstract

**Background:** Current clinical tests for mycobacterial pulmonary diseases (MPD), such as pulmonary tuberculosis (PTB) and non-tuberculous mycobacteria pulmonary diseases (NTM-PD), are inaccurate, time-consuming, sputum-dependent, and/or costly. We aimed to develop a simple, rapid and accurate breath test for screening and differential diagnosis of MPD patients in clinical settings.

**Methods:** Exhaled breath samples were collected from 93 PTB, 68 NTM-PD and 4 PTB&NTM-PD patients, 93 patients with other pulmonary diseases (OPD) and 181 healthy controls (HC), and tested using the online high-pressure photon ionisation time-of-flight mass spectrometer (HPPI-TOF-MS). Machine learning models were trained and blindly tested for the detection of MPD, PTB, NTM-PD, and the discrimination between PTB and NTM-PD, respectively. Diagnostic performance was evaluated by metrics of sensitivity, specificity, accuracy, and area under the receiver operating characteristic curve (AUC).

**Results:** The breath PTB detection model achieved a sensitivity of 73.5%, a specificity of 85.8%, an accuracy of 82.9%, and an AUC of 0.895 in the blinded test set (n=141). The corresponding metrics for the NTM-PD detection model were 86.4%, 93.2%, 92.1% and 0.972, respectively. For distinguishing PTB from NTM-PD, the model also achieved good performance with sensitivity, specificity, accuracy, and AUC of 85.3%, 81.8%, 83.9% and 0.947, respectively. 22 potential breath biomarkers associated with MPD were putatively identified and discussed, which included 2-furanmethanol, ethanol, 2-butanone, etc.

**Conclusions:** The developed breathomics-based MPD detection method was demonstrated for the first time with good performance for potential screening and diagnosis of PTB and NTM-PD using a refined operating procedure on the HPPI-TOF-MS platform.

## Introduction

Tuberculosis (TB) is a chronic infection and one of the top lethal diseases worldwide. Currently affecting one quarter of the global population, TB continues to pose a major health threat, with 10.6 million new cases estimated in 2021 by WHO [1]. Meanwhile, non-tuberculous mycobacteria (NTM) pulmonary diseases (PD) (NTM-PD) have recently seen a surge in cases, sharing similar symptoms, radiographic findings and pathological characteristics with pulmonary TB (PTB), leading to frequent misdiagnoses without strain identification [2]. Additionally, most NTMs possess natural resistance to anti-TB drugs, with therapeutic schemes different from TB.

Current tests for PTB and NTM-PD, including rapid molecular tests, etiological examinations, immunodiagnostic tests and chest radiography, are limited by their expenses, inaccuracy, time-consumption, low-compliance, sputum-dependence, invasiveness or complexity [3]. Therefore, it is crucial to develop rapid, easy-to-operate, accurate and sputum-free diagnostic methods in clinical practice of mycobacterial PD (MPD) management and control.

Various studies have demonstrated that specific volatile organic compounds (VOCs) were emitted from cultured TB and NTM bacteria [4], and detecting these characteristic VOCs might provide new insight into MPD diagnosis. Breath VOC tests have enabled the detection of many human infectious diseases [5] [6] as these VOCs are associated with the metabolites in body fluids and tissues, offering a simple and non-invasive window into MPDs such as PTB and NTM-PD.

Several studies have investigated TB screening and diagnosis with breath tests in humans [5] [7]. Phillips et al. demonstrated the feasibility of breath tests for PTB detection with 82.6% sensitivity and 100% specificity [8], and 85% accuracy in a transcontinental and ethnic group with gas chromatography-mass spectrometer (GC-MS) in PTB screening [9]. Subsequent investigations have revealed more potential TB breath VOC biomarkers [10] [11] [12] [13]. Recently, Beccaria et al. used GC×GC-MS to analyse exhaled VOCs of PTB patients and suspected PTB patients / other controls in South Africa / Haiti, achieving high sensitivity and specificity based on 23 / 22 feature VOCs [14] [15], respectively. Other recent studies have focused on pediatric [16] and adult PTB diagnosis [17] and TB infection (TBI) screening [18]. In addition to the above TB VOC-identified studies, sensor-based breath analyses have also shown promise in detecting PTB without biomarkers revealed [7].

Regarding breath NTM detection, only one pilot study has been conducted thus far. Mani-Varnosfaderani et al. [19] discovered 17 markers associated with NTM-PD and demonstrated that exhaled VOCs could differentiate active NTM-PD from the patients with indolent infection or no cultured NTM. However, this study only included 11 subjects. No breath research has differentiated NTM-PD from PTB or other pulmonary diseases (OPD) as yet.

In this study, for the first time, we investigated the potential of breath analysis to differentiate among PTB, NTM-PD, OPD and healthy controls (HC), and identified 22 potential VOC biomarkers associated with PTB and NTM-PD, extending our previous PTB breath detection research [17] [18]. Moreover, by employing our self-developed on-line mass spectrometer for human breath—the high-pressure photon ionisation time-of-flight mass spectrometry (HPPI-TOF-MS) [20], we built a rapid and simple online breath analysis and modeling method for MPD screening and diagnosis.

## Methods

### Study design and population

The study was conducted from September 1 to December 31, 2022, at Guangzhou Chest Hospital, China, with approval from the Ethics Committee of Guangzhou Chest Hospital (No. 2022-65). Written consent was obtained from all participants.

Illustrated in Fig. 1, subjects were breath-sampled and clinically assessed before being categorized into five groups: PTB, NTM-PD, PTB&NTM-PD, OPD, and HC. The data was randomly split for machine learning (ML) model training, validation and testing. Various ML models were trained based on multiple biomarkers, validated and finally evaluated on a blinded test set.

**FIGURE 1.**
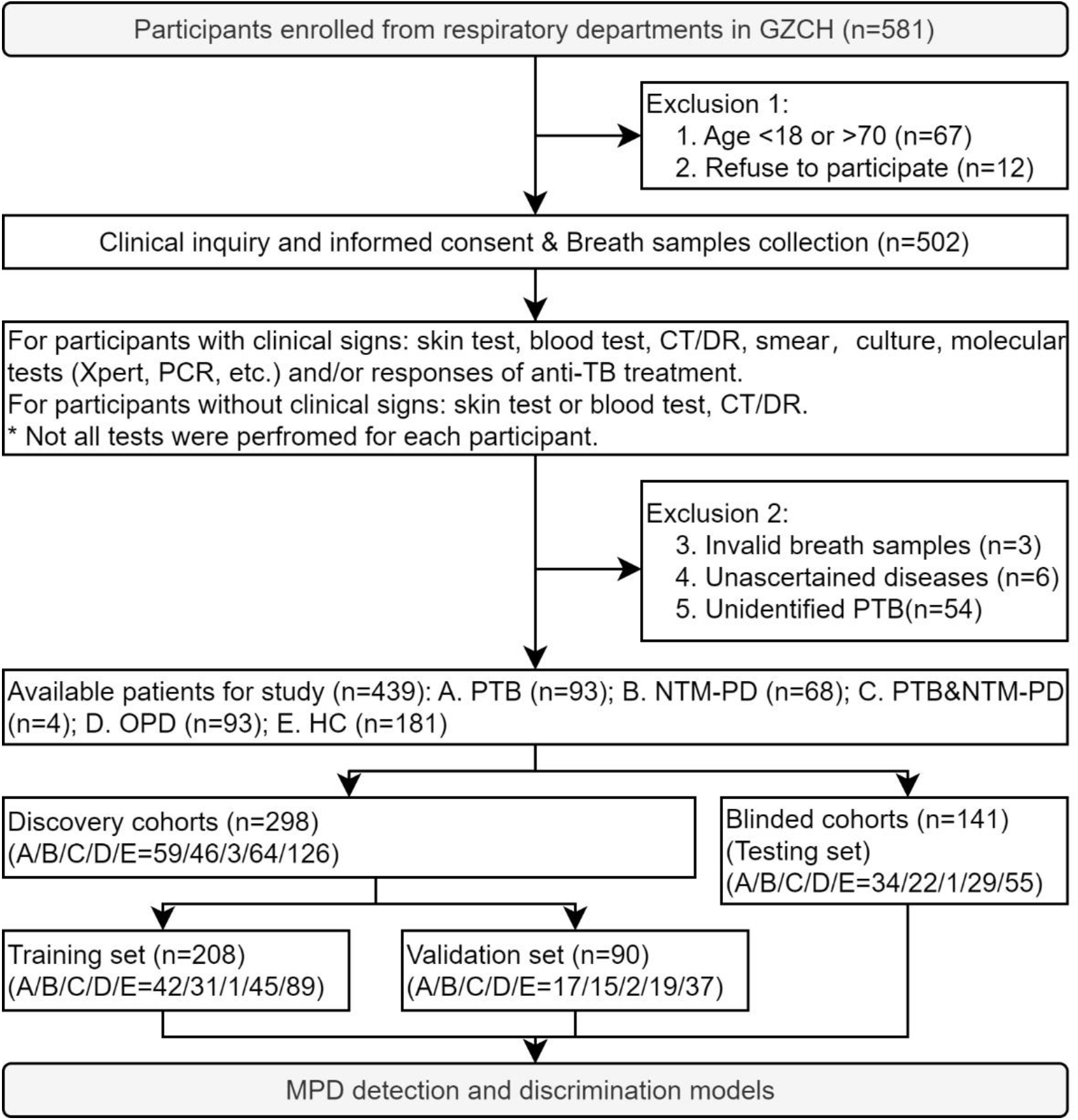
Enrollment of subjects and data division for model construction and test. GZCH: Guangzhou Chest Hospital (China), CT: computed tomography, DR: digital radiography, PCR: polymerase chain reaction, TB: tuberculosis, PTB: pulmonary TB, NTM-PD: non-tuberculous mycobacteria pulmonary diseases, OPD: other pulmonary diseases, HC: health controls, MPD: mycobacterial pulmonary diseases.

PTB and NTM-PD patients were prospectively recruited based on the following criteria: 1) diagnosed by Xpert, culture, smear, and/or other molecular tests; 2) before anti-TB/NTM treatment; 3) absence of OPDs. OPD patients were included if they had other pulmonary infectious diseases or noninfectious chronic diseases that shared similar symptoms with PTB or NTM-PD, such as pneumonia, lung cancer, lung abscess, etc. HCs had no respiratory symptoms and no pulmonary lesions on chest imaging. Note some HCs and OPDs might have TBIs. All participants were aged 18-70 years and the participants were excluded if their breath airbags leaked or if they were unable to inhale sufficient air (<1.2L).

### Breath sampling

The method has been detailed in our previous breath detection studies [17]. Briefly, breath samples were collected according to a predefined protocol and analysed within 24 hours. The sampling apparatus comprised a disposable gas nipple and a polyether-ether-ketone bag. Dietary and environmental factors were minimised through established sampling requirements, abstinence protocols and a consistent collection environment, including abstaining from smoking and alcohol before the sampling day, fasting for > 2 h and rinsing their mouths before sampling. Participants inhaled deeply and exhaled completely into the sampling bag with a volume ≥ 1.2 L.

### HPPI-TOF-MS analysis

The basic development and structure of HPPI-TOF-MS has been described in our previous study [20] with universal detection operations for breath VOCs [21]. Briefly, the device comprises a vacuum ultraviolet lamp-based HPPI ion source and an orthogonal acceleration TOF mass analyser.

The gas-phase exhaled breath sample was directly introduced into the 60 °C heated ionisation region through a 100 °C heated stainless-steel capillary from the air bag to eliminate condensation and adsorption of exhaled VOCs. Then, TOF signals were recorded with a 400-picosecond time-to-digital converter rate at 25 kHz and accumulated all the mass spectra for 60s. Finally, the bundled analysis software calibrated all mass spectrometers and completed noise reducing, baseline correction and background air subtraction before further analysis.

### Data analysis

As depicted in Fig. 1, participants were randomly divided into two groups: 70% for model construction and 30% for blinded testing. Given that the enroled datasets was discrete, balanced, and with a median feature scale, a multi-strategy feature selection approach was performed before the model construction to identify the most important VOC features for modeling. Firstly, the VOC ions with no significant differences (p > 0.05) between case and control groups were excluded. Then, the VOC ions with low intensity but highly correlated with other selected VOC ions (correlation coefficient > 0.9) among all training samples were excluded. Lastly, a random forest (RF) model was constructed on the training data, and the ten most important VOC ions were selected based on the feature importance or coefficient. Based on the finally selected VOC features, several well-established ML models were employed as classifiers to distinguish MPD patients from controls. These algorithms included RF, logistic regression (LR), extreme gradient boosting (XGB), k-nearest neighbours (KNN), decision tree (DT) and ensemble learning. Subsequently, the optimal classifier was selected according to the model performance in the internal validation subset, and tested with the receiver operating characteristic (ROC) curve analysis. Sensitivity (SEN), specificity (SPE), accuracy (ACC), area under the ROC curve (AUC) and their relative 95% confidence interval (CI) were calculated to evaluate the performance of MPD detection models on each top characteristic VOC ion and on all featured VOC ions in a panel. Brief descriptions and main parameters of all ML methods were provided in Table S1.

### Statistical analysis

All statistical analyses were conducted using the software IBM SPSS Statistics v24 (IBM Corp., Armonk, NY, USA) and Origin v2018 (OriginLab Corp., Northampton, MA, USA). Descriptive statistics are reported as frequencies (percentages) for categorical data and as medians (interquartile range, IQR) for continuous variables. The Mann-Whitney U test was used to compare the demographic characteristics between different patient groups for continuous variables and the chi-square test for categorical variables. All tests were two-tailed.

## Results

### Study population

We recruited 581 participants from multiple respiratory departments for PTB and NTM-PD in Guangzhou Chest Hospital. After excluding participants aged outside 18∼70, and those who refused the study or had invalid breath samples, 439 participants were finally eligible for analysis. Of these individuals, 93 were diagnosed with PTB (confirmed by aetiological tests), 68 with NTM-PD, 4 with both aetiological PTB and NTM-PD, 93 with OPD, and 181 HCs were finally enroled. The demographics of participants and their clinical information are presented in Table 1. Subjects with PTB and NTM-PD were diagnosed based on treatment responses or aetiological tests such as sputum smear, culture, and/or molecular tests including Xpert, TB/NTM-RNA, TB-LAMP and TB/NTM-PCR tests. 54 of all PTB patients were ultimately confirmed by clinical comprehensive diagnosis including 5 PTB&NTM-PD and thus excluded provisionally, suggesting the current aetiological tests are not entirely satisfactory. Most OPD participants had aetiological test results to exclude the comorbidity of PTB/NTM-PD, and HCs underwent immunological tests and chest radiography examinations for inclusion.

**TABLE 1.**
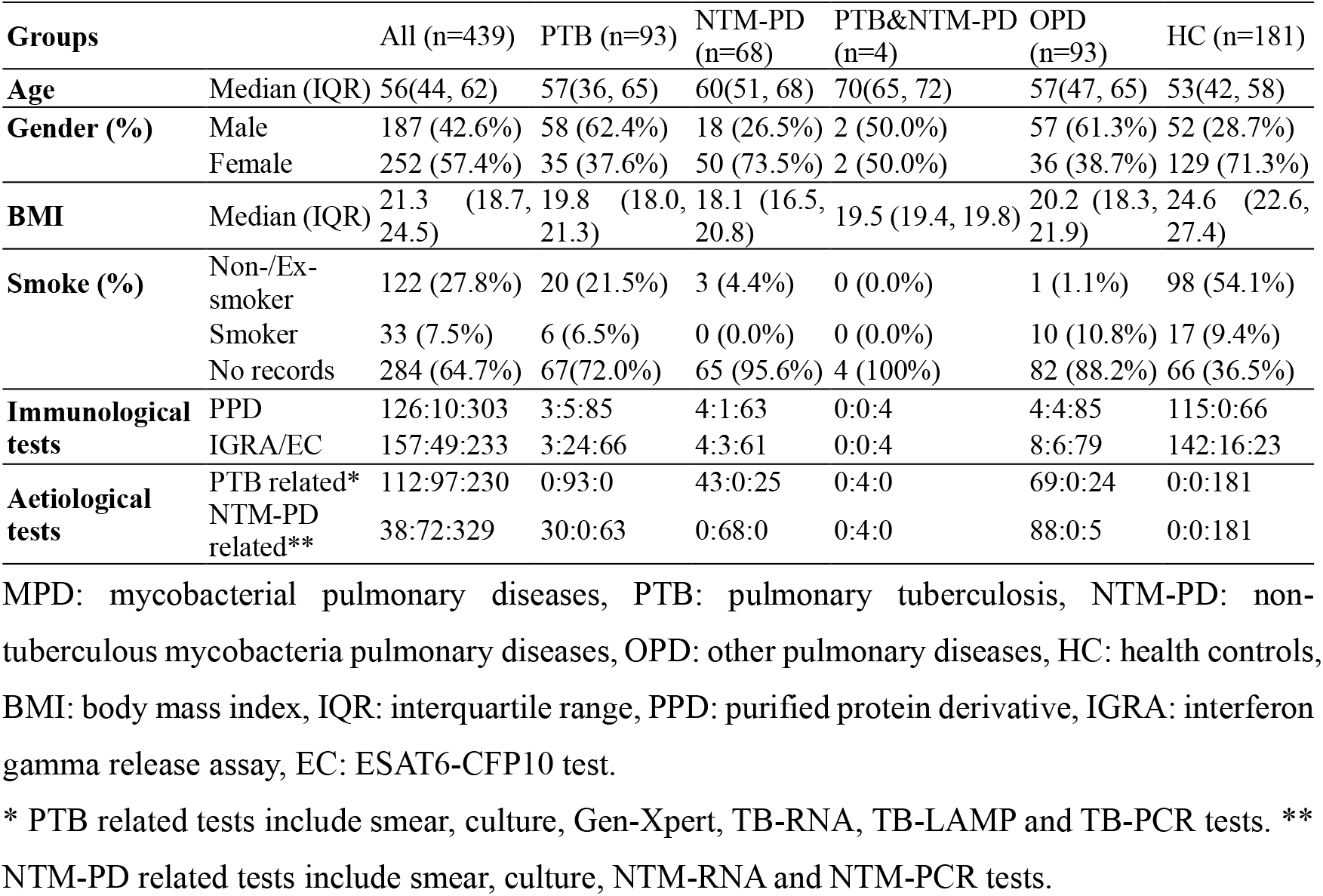
Basic demographic data of all enrolled participants and their MPD-related clinical test results (negative: positive: untested)

Statistical analyses were performed on the basic demographic characteristics of age, gender, body mass index (BMI) and smoking status between case and control groups for four tasks: MPD detection, PTB detection, NTM-PD detection and PTB&NTM-PD discrimination. As shown in Table S2, there were no significant differences in gender and smoking status between MPD group and controls, but with significant differences in age and BMI. For the PTB detection dataset, BMI and smoking status were significantly different between cases and controls. The greatest bias was observed between NTM-PD group and controls, with significant differences in all four characteristics. Although there was no significant difference in smoking status between PTB and NTM-PD, age, gender and BMI were all significantly biased for PTB&NTM-PD discrimination. In addition, the value of the statistical analysis results for smoking status were limited due to incomplete data collection.

### MPD detection performance

Firstly, we explored the potential value of breath VOC test in MPD detection. The classification performance metrics of six ML models for MPD detection were evaluated using a validation dataset, presented in Table S3. Among these results, RF, XGB and ensemble models achieved superior performances than other models, with AUC greater than 0.85. Overall, the ensemble model has the best performance in the validation set with a SEN of 76.5% (95% CI: 62.2%, 90.7%), a SPE of 89.3% (95% CI: 81.2%, 97.4%) and an AUC of 0.909 (95% CI: 0.849, 0.968). Therefore, the ensemble algorithm was further analysed in the testing set regarding different controls as shown in Table 2, and also employed as the basic classifier for other three tasks.

**TABLE 2.** The performance metrics of ensemble model on the test set in MPD, PTB, NTM-PD detection and PTBB&NTM-PD discrimination (95% CI).

| Models | Controls | SEN(%) | SPE(%) | ACC(%) | AUC |
| --- | --- | --- | --- | --- | --- |
| **MPD detection** | OPD | 73.7(62.3, 85.1) | 65.5(48.2, 82.8) | 70.9(61.3, 80.5) | 0.817(0.735, 0.898) |
|  | HC | 73.7(62.3, 85.1) | 94.5(88.5, 100) | 83.9(77.1, 90.7) | 0.946(0.904, 0.988) |
|  | All | 73.7(62.3, 85.1) | 84.5(76.8, 92.3) | 80.1(73.6, 86.7) | 0.901(0.852, 0.951) |
| **PTB detection** | NTM-PD | 73.5(58.7, 88.4) | 72.7(54.1, 91.3) | 73.2(61.6, 84.8) | 0.816(0.714, 0.917) |
|  | OPD | 73.5(58.7, 88.4) | 82.8(69.0, 96.5) | 77.8(67.5, 88.0) | 0.862(0.777, 0.947) |
|  | HC | 73.5(58.7, 88.4) | 92.7(85.9, 99.6) | 85.4(78.1, 92.7) | 0.944(0.897, 0.992) |
|  | All | 73.5(58.7, 88.4) | 85.8(79.2, 92.5) | 82.9(76.6, 89.1) | 0.895(0.844, 0.946) |
| **NTM-PD detection** | PTB | 86.4(72.0, 100) | 91.2(81.6, 100) | 89.3(81.2, 97.4) | 0.965(0.917, 1.000) |
|  | OPD | 86.4(72.0, 100) | 89.7(78.6, 100) | 88.2(79.4, 97.1) | 0.953(0.895, 1.000) |
|  | HC | 86.4(72.0, 100) | 96.4(91.4, 100) | 93.5(88.0, 99.0) | 0.986(0.960, 1.000) |
|  | All | 86.4(72.0, 100) | 93.2(88.7, 97.8) | 92.1(87.7, 96.6) | 0.972(0.944, 0.999) |
| **PTB vs NTM-PD** | NTM-PD | 85.3(73.4, 97.2) | 81.8(65.7, 97.9) | 83.9(74.3, 93.5) | 0.947(0.888, 1.000) |

MPD: mycobacterial pulmonary diseases, PTB: pulmonary tuberculosis, NTM-PD: non-tuberculous mycobacteria pulmonary diseases, OPD: other pulmonary diseases, HC: healthy controls, SEN: Sensitivity, SPE: Specificity, ACC: accuracy, AUC: area under the receiver operating characteristic curve. CI: confidence intervals.

From Table 2, it can be seen that the ensemble based MPD detection model achieved higher SPE and AUC of 94.5% and 0.946 in discriminating MPD from HC than those in distinguishing MPD from OPD (65.5% and 0.817), respectively. This may suggest that the metabolic differences between MPD and HC were larger than those between MPD and OPD. Overall, the MPD detection model achieved a SEN of 73.7% (95% CI: 62.3%, 85.1%), a SPE of 84.5% (95% CI: 76.8%, 92.3%) and an AUC of 0.901 (95% CI: 0.852, 0.951) in discriminating MPD and all controls in clinical practice. The corresponding ROC curves for MPD detection model is illustrated in Fig. 2 (a), which includes MPD recognition from all controls, OPD and HC in validation and test sets.

**FIGURE 2.**
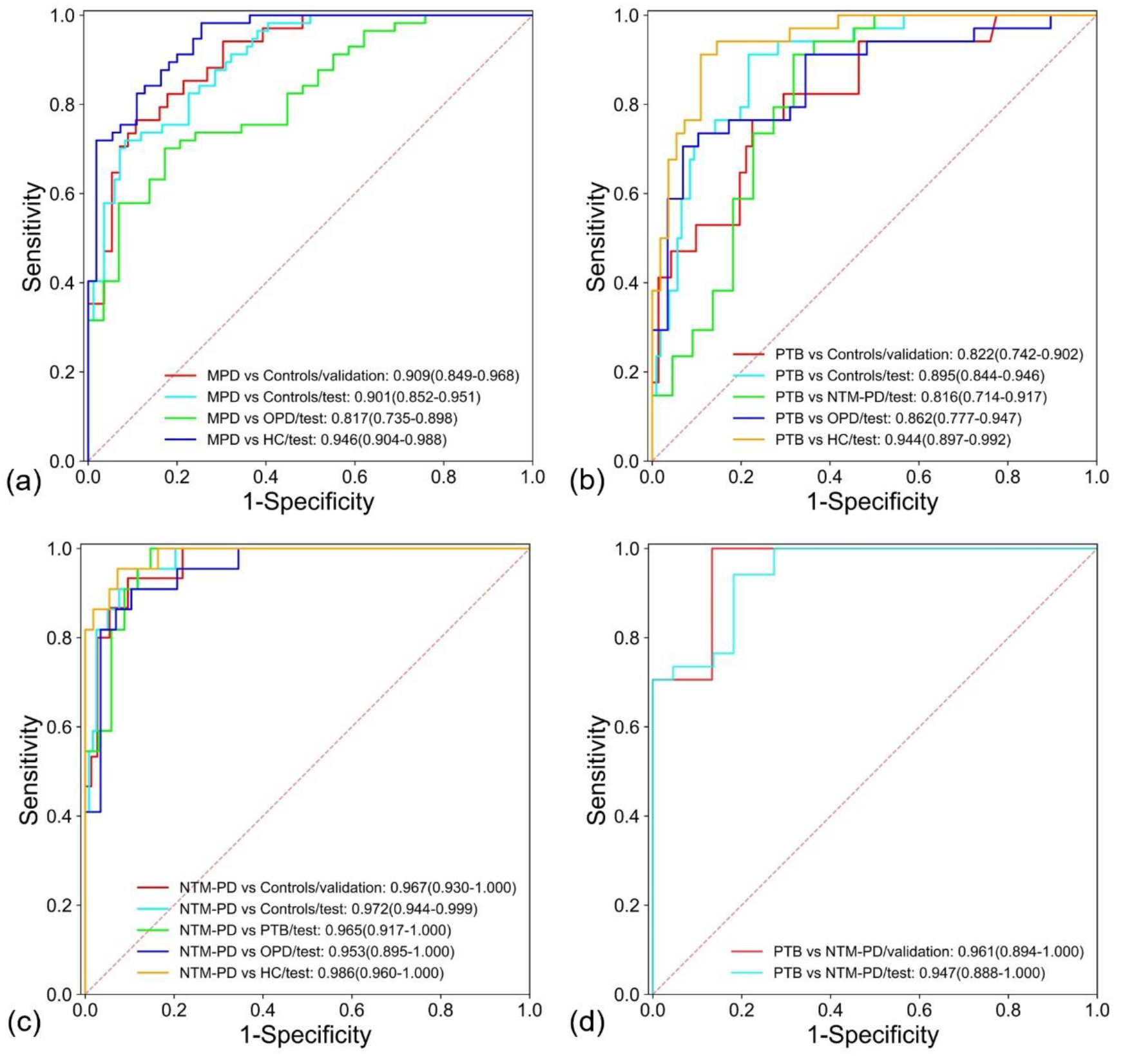
The performance of ensemble based MPD, PTB, and NTP-PD detection models, and PTB&NTM-PD discrimination model. (a) ROC curves of MPD detection model in validation and test sets. (b) ROC curves of PTB detection model in validation and test sets. (c) ROC curves of NTM-PD detection model in validation and test sets. (d) ROC curves of PTB&NTM-PD discrimination model in validation and test sets. MPD: mycobacterial pulmonary diseases, PTB: pulmonary tuberculosis, NTM-PD: non-tuberculous mycobacteria pulmonary diseases, ROC: receiver operating characteristic, OPD: other pulmonary diseases, HC: health controls.

### PTB, NTM-PD detection and PTB&NTM-PD discrimination performances

Based on similar feature selection and modeling processes, a PTB detection model based on ensemble method was trained and blindly tested, with performance metrics presented in Table 2. The AUCs for PTB discrimination from all control or different subgroups of controls were all greater than 0.8 (range: 0.816∼0.944). These results indicate that PTB can be distinguished with other symptomatically similar diseases, such as NTM-PD and other common lung diseases. Fig. 2 (b) illustrates the ROC curves of PTB detection model for distinguishing PTB from NTM-PD, OPD and HC.

Similarly, a NTM-PD detection model was also developed and blindly tested. As shown in Fig. 2 (c), the AUCs of NTM-PD discrimination from other groups using the panel of characteristic VOCs were more satisfactory, ranging from 0.953 to 0.986, suggesting that NTM-PD could be easily distinguished in complex clinical settings through breath VOC analysis.

Additionally, the performance of a diagnosis model for discriminating PTB from NTM-PD was shown in Fig. 2 (d) and Table 2. The SEN, SPE, ACC and AUC were 85.3% (95% CI: 73.4%, 97.2%), 81.8% (95%CI: 65.7%, 97.9%), 83.9% (95% CI: 74.3%, 93.5%) and 0.947 (95% CI: 0.888, 1.000), respectively.

As an extension, we additionally evaluated the performance of aforementioned models on the 49 excluded clinically diagnosed PTB patients who had negative aetiological TB and NTM results. As illustrated in Fig. S1, these models, especially the NTM-PD detection and PTB&NTM-PD discrimination models, could tentatively assign these patients with elevated probabilities of PTB.

### Potential VOC biomarkers related to MPD

Based on the top ten VOCs employed by the above MPD, PTB and NTM-PD detection and PTB&NTN-PD discrimination tasks, 22 VOC ions were identified as potential biomarkers associated with MPD (one was identified as fragments of carboxylic acids/esters). To evaluate the discrimination power of these VOC biomarkers, we trained the detection models in the corresponding tasks on each single VOC ion and evaluated them in the test dataset. As illustrated in Fig. 3 (a), the classification AUCs of each VOC ions ranged from 0.40 to 0.92 in all tasks, which were all inferior to that of the panel of multiple VOC ions in corresponding tasks. There is one VOC ion (m/z=99) selected in all four tasks, four VOC ions (m/z=45, 47, 55 and 73) selected in three tasks, seven VOC ions (m/z=78, 67, 69, 70, 115, 81 and 106) selected in two tasks, and ten other VOC ions selected in only one task. Thus, we ranked the 22 VOC ions by the probability of being selected in four tasks as in Fig. 3 (b), where the probability represents the importance of the corresponding VOC ion as a biomarker of MPD. The relative concentration (peak area) of all the ranked 22 VOC ions in HC, OPD, NTM-PD, and PTB groups was illustrated in Fig. 4. The connection line indicates a significant difference between the two groups being connected. It can be observed that, (1) there is significant differences between at least two groups in all selected VOC ions; (2) there are significant differences between any two groups in three VOC ions (m/z=55, 99 and 72); (3) there are significant differences between PTB and NTM-PD in all VOC ions except the icons with m/z 106, 83, 105 and 91.

**FIGURE 3.**
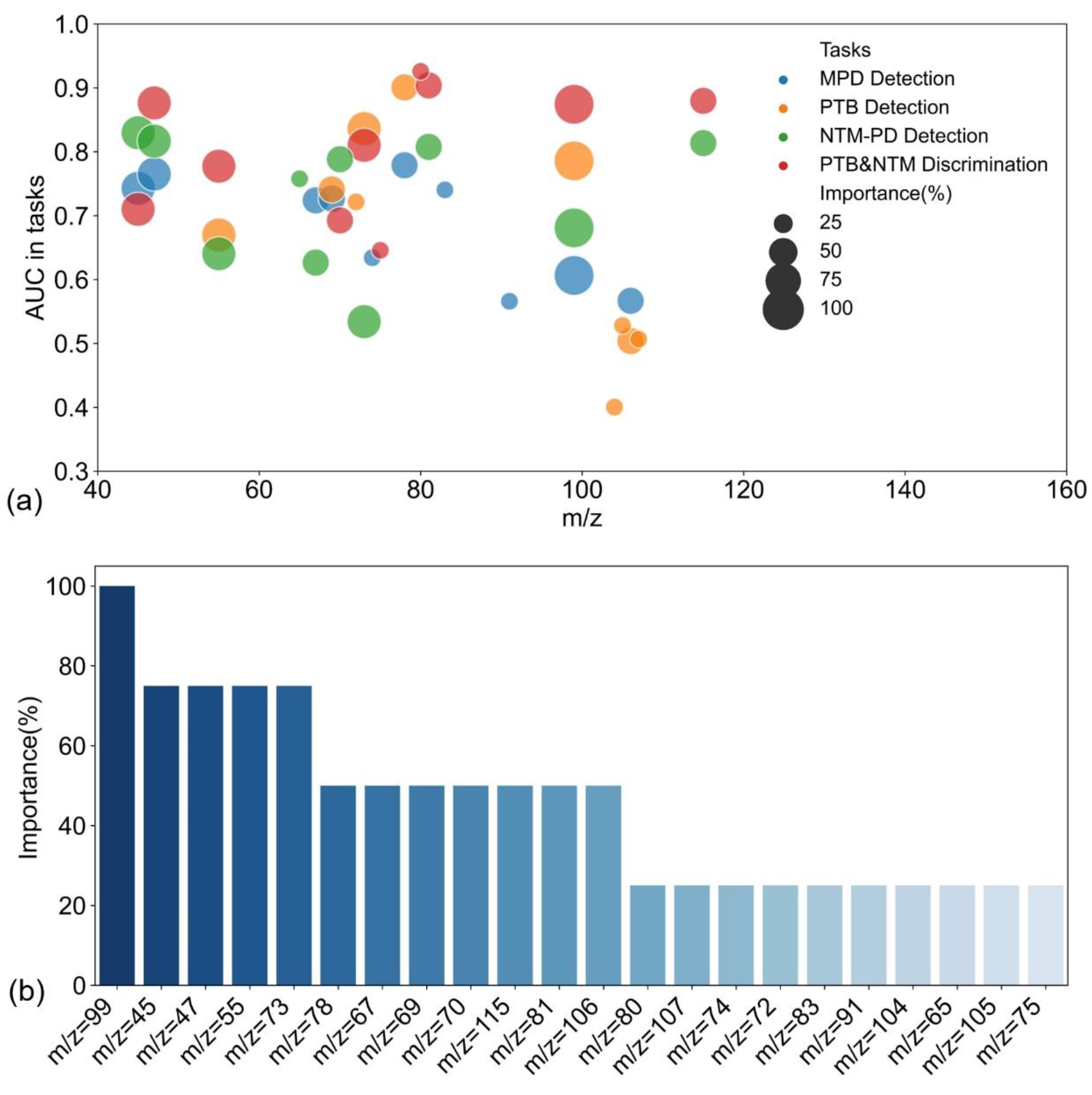
The characteristics analysis of the 22 potential VOC biomarker ions identified in this study. (a) The discrimination power and m/z of the 22 VOC ions by bubble chart of involved tasks (bubble color) and the probability of being selected in four tasks (bubble size). (b) The ranked VOC ions based on the probability of being selected in four tasks. VOC: volatile organic compounds, MPD: mycobacterial pulmonary diseases, PTB: pulmonary tuberculosis, NTM-PD: non-tuberculous mycobacteria pulmonary diseases, AUC: area under the receiver operating characteristic curve.

**FIGURE 4.**
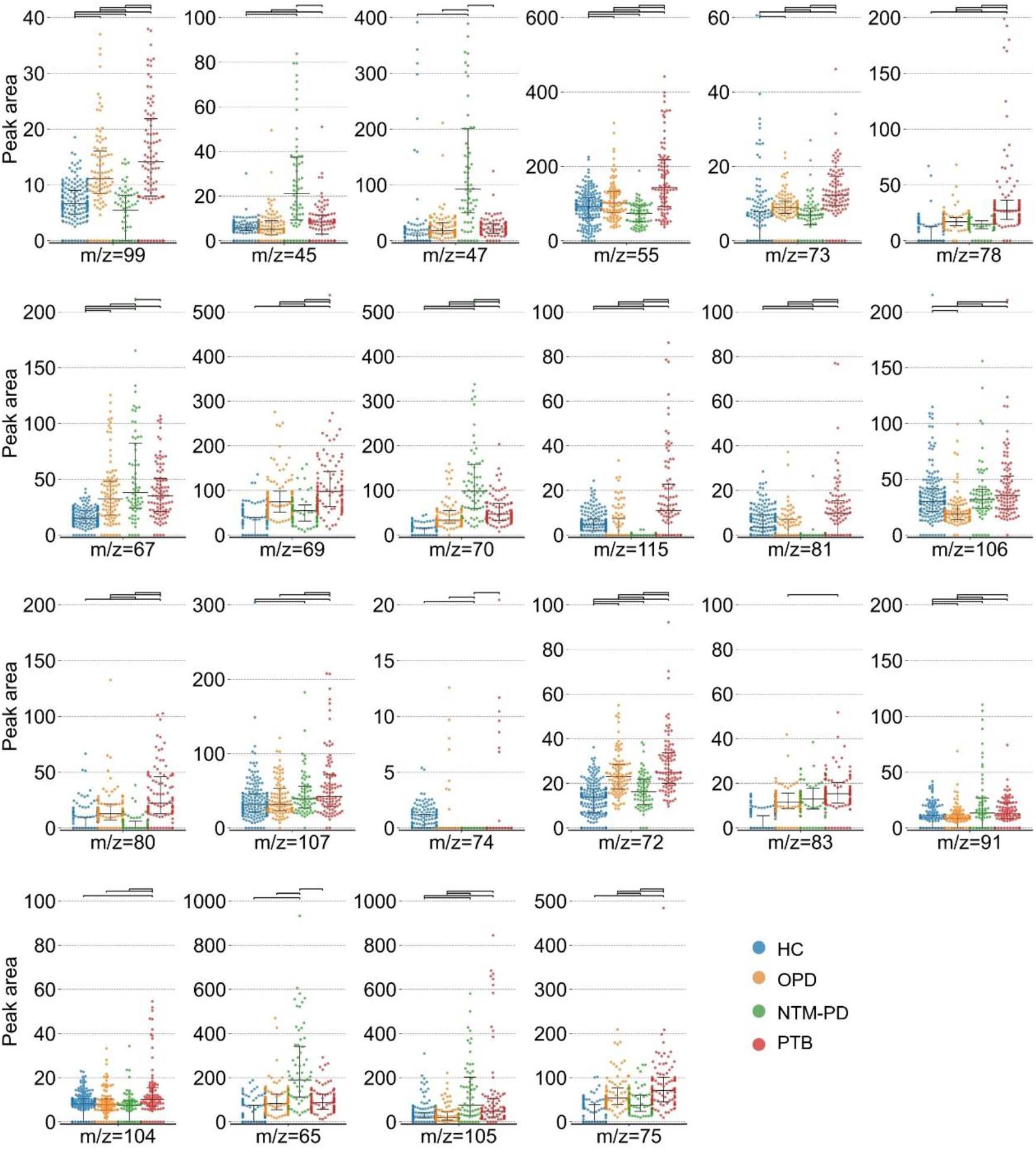
Relative concentrations of the 22 potential VOC biomarker ions ranked by probability detected in OPD, NTM-PD and PTB groups. The connection line indicates a significant difference between the two groups being connected. VOC: volatile organic compounds, MPD: mycobacterial pulmonary diseases, PTB: pulmonary tuberculosis, NTM-PD: non-tuberculous mycobacteria pulmonary diseases, HC: health controls.

Given that the TOF-MS can only confirm the m/z of detected VOCs, the possible chemical identities of these ions were inferred based on the peak area distribution in addition to m/z data, comparison with published potential VOC biomarkers for TB and NTM and the human breathomics database [22]. These VOC ions selected in at least two tasks (importance≥50%) were tentatively assigned as furfuryl alcohol (m/z=99, CAS number: 98-00-0), fragments of carboxylic acids/esters (m/z=45), ethanol (m/z=47, 64-17-5), 2-cyanoethyl radical (m/z=55, 25840-11-3), butanal/2-butanone (m/z=73, 123-72-8/78-93-3), Benzene (m/z=78, 71-43-2), 3-butenenitrile (m/z=67, 109-75-1), isobutyronitrile (m/z=69, 78-82-0), 1-pentene (m/z=70, 109-67-1), N-isobutylacetamide (m/z=115, 1540-94-9), 1-methylpyrrole (m/z=81, 96-54-8) and m-Xylene/benzaldehyde (m/z=106, 108-38-3/100-52-7), among the full list of 22 potential VOC biomarkers detailed in Table 3.

**TABLE 3.** The key VOCs associated with MPD according to selected probability ≥25% in all four tasks.

| No. | m/z | Related tasks* | Potential VOCs | CAS number | Molecular weight | Molecular formula |
| --- | --- | --- | --- | --- | --- | --- |
| 1 | 99 | A, B, C, D | Furfuryl alcohol | 98-00-0 | 98.100 | C<sub>5</sub>H<sub>6</sub>O<sub>2</sub>(+H) |
| 2 | 45 | A, C, D | Fragments of carboxylic acids/esters | / | / | COOH<sup>+</sup> |
| 3 | 47 | A, C, D | Ethanol | 64-17-5 | 46.068 | C<sub>2</sub>H<sub>6</sub>O(+H) |
| 4 | 55 | B, C, D | 2-Cyanoethyl radical | 25840-11-3 | 54.071 | C<sub>3</sub>H<sub>4</sub>N(+H) |
| 5 | 73 | B, C, D | Butanal/2-Butanone | 123-72-8/78-93-3 | 72.106 | C<sub>4</sub>H<sub>8</sub>O(+H) |
| 6 | 78 | A, B | Benzene | 71-43-2 | 78.112 | C<sub>6</sub>H<sub>6</sub> |
| 7 | 67 | A, C | 3-Butenenitrile | 109-75-1 | 67.089 | C<sub>4</sub>H<sub>5</sub>N |
| 8 | 69 | A, B | Isobutyronitrile | 78-82-0 | 69.105 | C<sub>4</sub>H<sub>7</sub>N |
| 9 | 70 | C, D | 1-Pentene | 109-67-1 | 70.133 | C<sub>5</sub>H<sub>10</sub> |
| 10 | 115 | C, D | N-Isobutylacetamide | 1540-94-9 | 115.174 | C<sub>6</sub>H<sub>13</sub>NO |
| 11 | 81 | C, D | 1-Methylpyrrole | 96-54-8 | 81.116 | C<sub>5</sub>H<sub>7</sub>N |
| 12 | 106 | A, B | m-Xylene/Benzaldehyde | 108-38-3/100-52-7 | 106.165/106.122 | C<sub>8</sub>H<sub>10</sub>/C<sub>7</sub>H<sub>6</sub>O |
| 13 | 80 | D | Pyridine | 110-86-1 | 79.100 | C<sub>5</sub>H<sub>5</sub>N(+H) |
| 14 | 107 | B | 2,6-Dimethylpyridine | 108-48-5 | 107.153 | C<sub>7</sub>H<sub>9</sub>N |
| 15 | 74 | A | Propionic acid | 79-09-4 | 74.078 | C<sub>3</sub>H<sub>7</sub>O<sub>2</sub> |
| 16 | 72 | B | Butanal/2-Butanone | 123-72-8/78-93-3 | 72.106 | C<sub>4</sub>H<sub>8</sub>O |
| 17 | 83 | A | Pentanenitrile | 110-59-8 | 83.132 | C<sub>5</sub>H<sub>9</sub>N |
| 18 | 91 | A | 2-Ethoxyethanol | 110-80-5 | 90.121 | C<sub>4</sub>H<sub>10</sub>O<sub>2</sub>(+H) |
| 19 | 104 | B | 2-Methyl-1-butanethiol/Pentanethiol | 187-18-8/110-66-7 | 104.214 | C<sub>5</sub>H<sub>12</sub>S |
| 20 | 65 | C | Cyanoallene | 1001-56-5 | 65.073 | C<sub>4</sub>H<sub>3</sub>N |
| 21 | 105 | B | Isopentanethiol | 541-31-1 | 104.214 | C<sub>5</sub>H<sub>12</sub>S(+H) |
| 22 | 75 | D | Glycine | 56-40-6 | 75.067 | C<sub>2</sub>H<sub>5</sub>NO<sub>2</sub> |

CAS: chemical abstracts service, VOCs: volatile organic compounds, MPD: mycobacterial pulmonary diseases.
\*Tasks: A. MPD detection; B. PTB detection, C. NTM-PD detection; D. PTB&NTM-PD discrimination.

## Discussion

The diagnosis of MPDs presents a significant challenge, particularly in the detection of PTB and NTM-PD. Current diagnostic methods for PTB and NTM-PD, including sputum culture, molecular tests and anti-TB or anti-NTM-PD treatments, are costly, time-consuming, and often inaccurate or inconvenient. Breathomics offers a novel approach to MPD detection through exhaled breath VOCs analysis, with promising results in PTB detection [5] [7]. In this study, we investigated the diagnostic value of breath analysis for MPD in a large cohort and successfully differentiated PTB and NTM-PD from the controls using breath VOCs for the first time. Models for MPD detection, PTB detection, NTM-PD detection and PTB&NTM-PD discrimination were trained and tested blindly with satisfactory SENs, SPEs, ACCs and AUCs, which demonstrated the potential diagnostic value of breath VOCs in clinical practice of MPD. Additionally, we putatively identified potential VOC biomarkers associated with MPD based on the most important model features, a review of the literature and breathomics databases, providing a foundation for the development of a breath analyser for PTB and NTM-PD in future comprehensive clinical practice.

For the NTM-PD detection, our study differs from the work of Mani-Varnosfaderani et al. [19], which pay attention to differentiating active NTM-PD from patients with indolent infection or no cultured NTM using breath analysis. We further focused on the differential diagnosis of NTM-PD from other symptom-like diseases including PTB and OPD, and HCs. Regarding PTB detection, we previously built a breath VOC model to differentiate 518 Mycobacterium TB (MTB) positive PTB patients from 810 HCs and/or 77 OPDs with high AUCs of 0.975 (PTB vs. Control) and 0.961 (PTB vs. OPD) and putatively identified 10 breath VOC biomarkers associated to PTB [17]. This study further extended this breathomics analysis to NTM-PD and MTB-negative PTB detection and more differential diagnosis tests. MTB-negative PTB patients with negative sputum culture and molecular test results, are commonly encountered in outpatient departments for TB and are often missed or misdiagnosed in practice. To the best of our knowledge, this is the first TB breath study to extend etiological sample trained models to these special patients (comprising about 1/3 of the whole enroled PTB group) and the elevated performance indicated that breath VOCs may provide a more precise diagnosis for the above challenge cases than traditional sputum-based tests.

Among the potential VOC biomarkers identified in this study, seven VOCs, namely ethanol, butanal, benzene, 2-butanone, 3-butenenitrile, isobutyronitrile and benzaldehyde, were found to be associated with PTB and/or NTM-PD. Ethanol has been reported as a VOC biomarker in the NTM detection study of Mani-Varnosfaderani et al. on human breath, in which a decreased concentration of ethanol was observed in the NTM-PD group [19]. In addition, Somashekar et al. [23] found that ethanolamine decreased in serum samples of guinea pigs infected with MTB after 30 days, and ethanolamine is easily synthesized by NH_3_and ethanol. Similarly, Ding et al. [24] observed a decrease of ethanol in the blood of both MTB infected human and mice, and the whole zebrafish larvae with M. marinum. However, an increase of ethanol was detected in PTB, NTM-PD and OPD groups in our study. Weber et al. also found the increase of ethanol in the breath of dairy cows infected with M. paratuberculosis [25] and de Laurentiis et al. showed that ethanol increased in the breath condensate of patients with chronic obstructive pulmonary disease (COPD) and pulmonary Langerhans cell histiocytosis [26]. Furthermore, ethanol was found in the headspace of cultures of M. paratuberculosis isolated from goat and cattle tissues and feces [27]. Spooner et al. detected widely varied levels of ethanol in serum of wild badgers with M. bovis [28]. Butanal was assigned as one of the characteristic breath VOCs in a case control study distinguishing mice infected with M. bovis BCG from the healthy [29]. Benzene was reported as a breath biomarker for PTB human by Beccaria et al. [15], and 2-butanone [8] [17] and 3-butenenitrile [17] have also been reported as breath biomarkers for PTB patients. Moreover, isobutyronitrile, benzene and 2-butanone have been identified as VOC markers in the headspace of MTB cultures [12] [30] and M. bovis BCG cultures [31]. Benzaldehyde has been associated with the breath [32] and feces [33] of cattle with M. bovis.

Our study has several strengths. First, breath testing, as a sputum-free diagnostic method, offers advantages in specimen sampling, especially for children and adults who have difficulty in producing sputum. Furthermore, we enroled the largest-ever cohort of MPD patients with PTB and NTM-PD in this pilot breath analysis. Third, we effectively differentiated NTM-PD from PTB and OPD with satisfactory SEN and SPE, and preliminarily extended our models in the clinically-challenging diagnosis of MTB-negative PTB patients. Finally, we employed the human breath mass spectrometer for rapid and convenient detection. While VOC detection has been applied in screening, diagnosis and treatment evaluation of numerous diseases, most studies have utilized either time-consuming and operationally complex GC-MS [34] or sensor-based electronic noses with limited receptive range and no qualitative or quantitative capability [35]. Both approaches have been proven difficult to translate to the clinical practice. In contrast, our study used the HPPI-TOF-MS for MPD, PTB and NTM-PD detections on breath samples. This instrument is designed for online fast breath sample analysis (1 min per sample), since it is equipped with an improved photon ionisation source and a high-pressure environment for lower detection limits (10 ppt) and high accuracy in human VOCs identification [20].

However, there are several limitations. Firstly, the potential biomarkers have not been completely confirmed. This is a common issue in VOC detection studies due to the early stage of this field and the limited capability of online MS. Further investigations and validations are needed to refine more consistent and precise panel VOCs for PTB and NTM-PD. Secondly, the metabolic pathways of the potential biomarkers are poorly understood. It is less clinically convincing for breath VOC diagnosis without clear origins and metabolic mechanisms of these molecules. Further basic biological and medical research are very necessary for the area of breathomics. Researchers have initiated explorations into the biosynthetic pathways of TB VOC markers using multi-omics and computational approaches [36], and links have been found between VOCs of breath of TB patients and VOCs in the headspace of MTB cultures [8] [12]. Thirdly, we have only completed the discrimination among general PTB, NTM-PD and OPD in adults. Further analysis will be conducted regarding subdivided groups, such as patients of MTB-negative PTB, MTB-positive PTB, NTM-PD with M. avium-intracellulare complex, NTM-PD with M. abscessus, etc. More pilot and validation studies are needed for breath detections of extra PTB, NTM infections outside the lung, children, people with comorbidities such as diabetes and HIV, etc. Finally, this is a single-centre pilot study, and there are some biases in the basic demography characteristics of enrolled participants. Multi-centre validations on independent cohorts are required for model and panel biomarker optimisation.

In conclusion, this study developed and evaluated a rapid and accurate breath test for the potential diagnosis of PTB and NTM-PD using online HPPI-TOF-MS. The results indicate that the proposed breathomics technology and method can discriminate PTB/NTM-PD from complex control group with good accuracy. Twenty-two potential VOC biomarkers were identified for MPDs screening and diagnosis. Further analysis on subdivided group differentiations and more extensive cohort studies are required before clinical application.

## Data Availability

All data produced in the present study are available upon reasonable request to the authors.

## Acknowledgements

We thank all the assistants, caregivers and enroled patients who participated in this study in PCAB Research Centre of Breath and Metabolism and Guangzhou Chest Hospital.

## Author contributors

Substantial contribution to conception and design: Y. Feng, J. Zhu, B. Su, H. Chen and Y. Tan. Data collection: L. Wu, Q. Sheng, P. Guan, P. Chen, H. Kuang, D. Li, W. Wang, Z. Feng, and Y. Tang. Statistical analysis: H. Chen and Y. Tan. Modeling and data interpretation: H. Chen and J. Zhu. Biomarker analysis: Y. Feng, M. He and H. Chen. Manuscript drafting: Y. Feng. Manuscript revising: H. Chen, Y. Feng, J. Zhu, B. Su and Y. Tan. The corresponding author, Y. Tan, had the final responsibility to submit the manuscript. All authors approved the submitted version of the manuscript.

---

**FIGURE S1.**
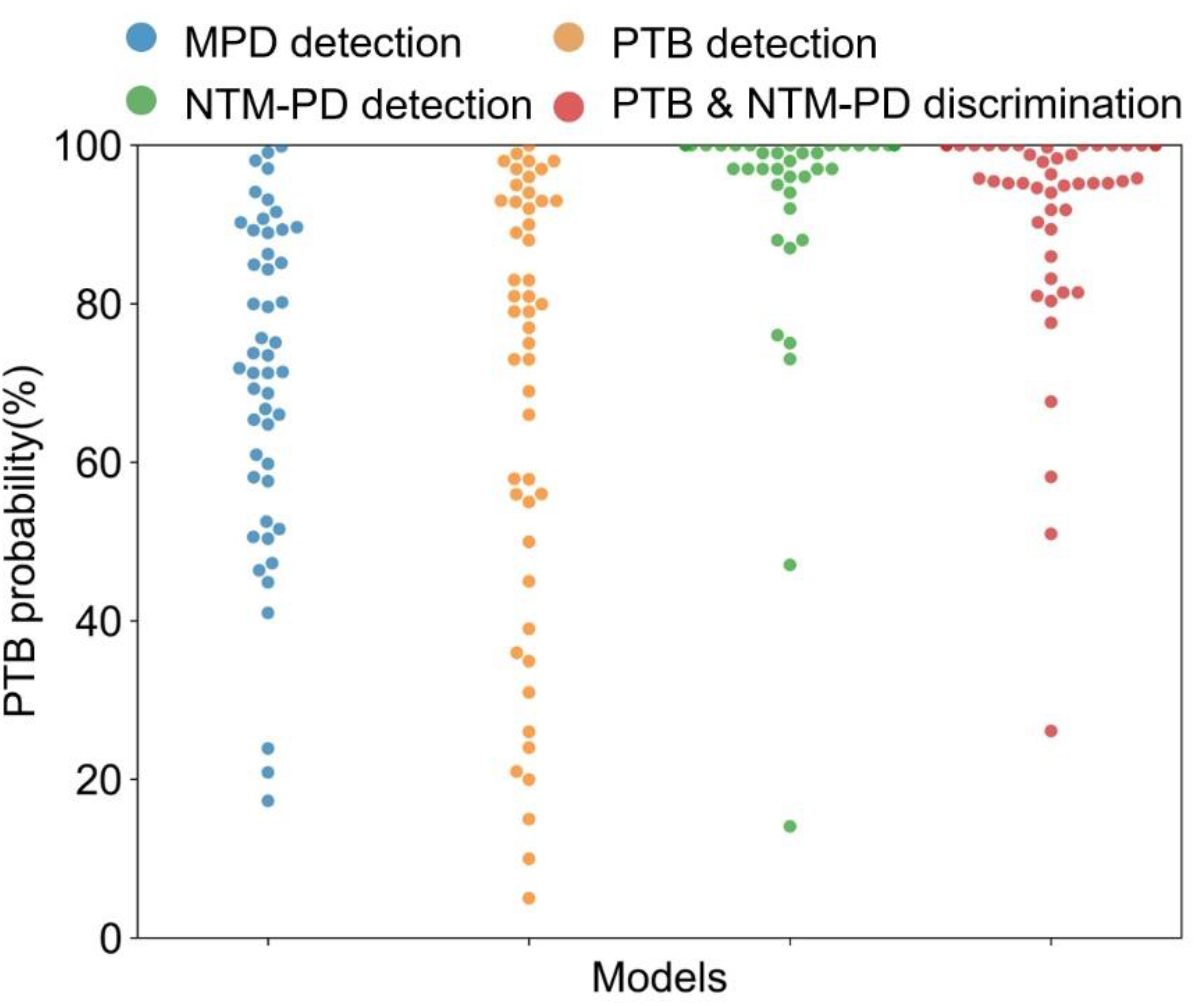
The predicted PTB probability comparison of the proposed models in 49 clinical confirmed PTB patients who were not confirmed by aetiological tests. MPD: mycobacterial pulmonary diseases, PTB: pulmonary tuberculosis, NTM-PD: non-tuberculous mycobacteria pulmonary diseases.

**TABLE S1.** The descriptions and main parameter settings of the employed ML models.

| **ML models** | **Descriptions** | **Main parameter settings\*** |
| --- | --- | --- |
| **RF** | A meta estimator that fits a number of decision tree classifiers on various sub-samples of the dataset and uses averaging to improve the predictive accuracy and control over-fitting. | n_estimators=100, max_features=0.5, min_samples_split=4, min_samples_leaf=10, criterion="entropy". |
| **LR** | It estimates the probability of an event occurring based on a given dataset of independent variables. | tol=1e-5, C=5.0, max_iter=1e+4. |
| **XGB** | A boosting algorithm based on gradient boosted decision trees. | booster: "gbtree", max_depth: 8, n_estimators: 100, min_child_weight: 3, gamma: 0.15, lambda: 2. |
| **KNN** | It achieves classification or prediction with the k-nearest neighbors voting. | n_neighbors=5, algorithm='auto', leaf_size=30, p=2, metric='minkowski'. |
| **DT** | It employs a divide and conquer strategy by conducting a greedy search to identify the optimal split points within a tree. | criterion="gini", splitter="best", min_samples_split=2, min_samples_leaf=1. |
| **Ensemble** | A voting classifier of RF, LR and XGB. | tol=1e-5, C=5.0, max_iter=10000, voting='soft'. |

ML: machine learning, RF: random forest, LR: logistic regression, XGB: extreme gradient boosting, KNN: k-nearest neighbors, DT: decision tree.
\* These algorithms were achieved based on python packages: xgboost
(https://xgboost.readthedocs.io/en/stable/python/python_intro.html) and sklearn (https://scikit-learn.org/stable/user_guide.html).

**TABLE S2.**
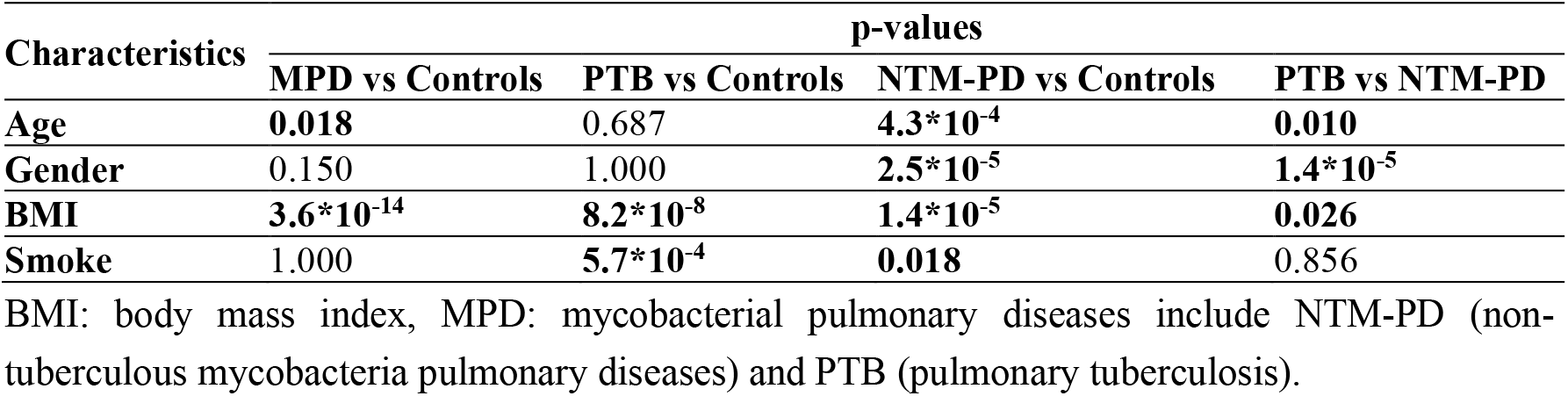
Statistical analysis of the basic demographic characteristics between the case and control groups in the MPD, PTB, and NTM-PD detection, and PTB&NTM-PD discrimination tasks.

| Characteristics | p-values |  |  |  |
| --- | --- | --- | --- | --- |
|  | MPD vs Controls | PTB vs Controls | NTM-PD vs Controls | PTB vs NTM-PD |
| Age | **0.018** | 0.687 | **$4.3 \times 10^{-4}$** | **0.010** |
| Gender | 0.150 | 1.000 | **$2.5 \times 10^{-5}$** | **$1.4 \times 10^{-5}$** |
| BMI | **$3.6 \times 10^{-14}$** | **$8.2 \times 10^{-8}$** | **$1.4 \times 10^{-5}$** | **0.026** |
| Smoke | 1.000 | **$5.7 \times 10^{-4}$** | **0.018** | 0.856 |

BMI: body mass index, MPD: mycobacterial pulmonary diseases include NTM-PD (non-tuberculous mycobacteria pulmonary diseases) and PTB (pulmonary tuberculosis).

**TABLE S3.**
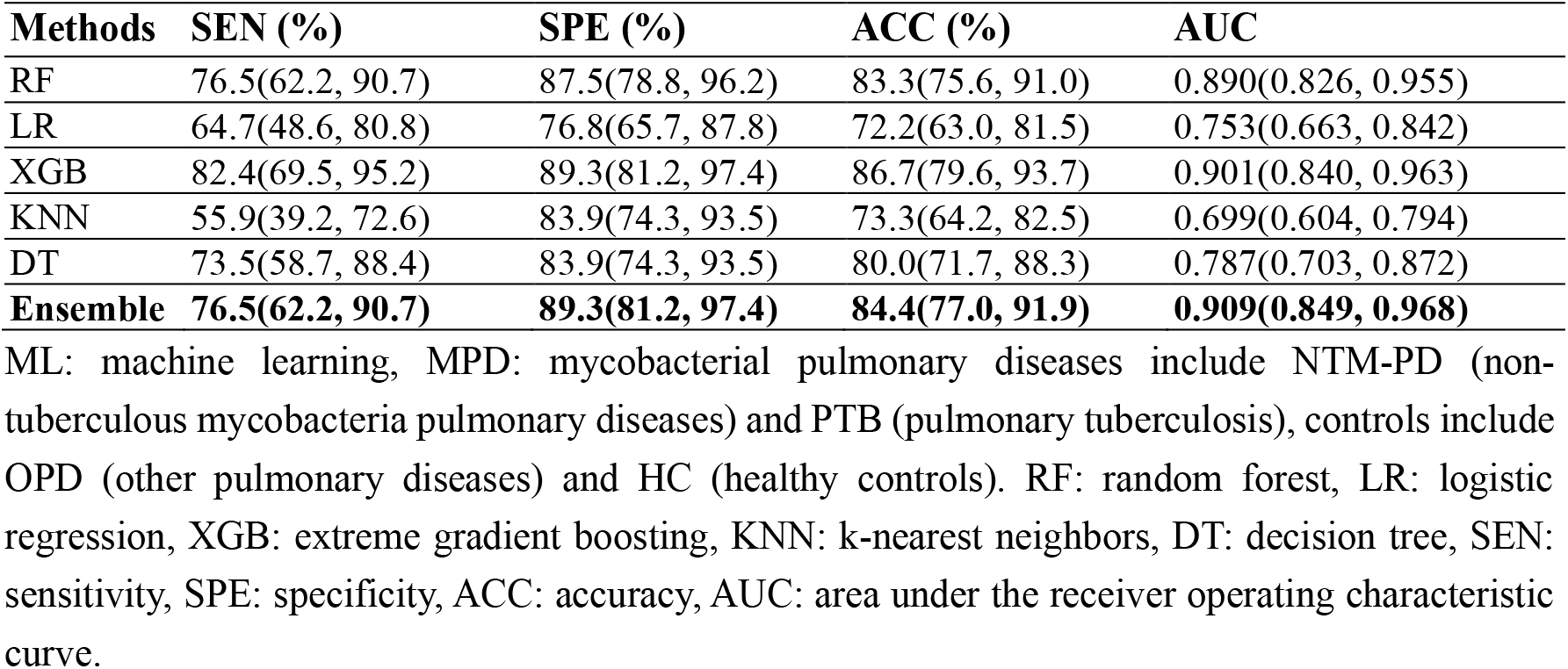
The performance metrics of six ML methods on the validation dataset in discriminating MPD from controls.

